# Estimated dietary intakes of vitamin A5

**DOI:** 10.1101/2024.10.07.24315069

**Authors:** Torsten Bohn, Marta Despotovic, Farhad Vahid, Ralph Rühl

## Abstract

A new vitamin concept, termed vitamin A5, was recently identified. Furthermore, dietary recommendations in the range of 1,1 (0,5 – 1,8) mg vitamin A5 / day were suggested by an international expert consortium. The ensuing question arises as to the current daily dietary intake amounts in the Western civilization. Addressing this answer included calculating the intake based on known amounts of vitamin A5 in frequently consumed food items of the human diet high in this vitamin as well as the known daily dietary intake amounts of those selected food components in Westernized countries. Regarding food items, amounts of vitamin A5 in the form of provitamin A5 (i.e. 9-*cis*-beta-carotene (9CBC)), the predominant form in the diet, were found to range from 0,1 to 39 µg 9CBC / g for individual fruits and vegetables, with highest concentrations in leafy vegetables. The average intake amounts of vitamin A5 in adults of the general population following a Western lifestyle in Europe, averaged 0,9 with a range of 0,5 (for Austria) – 1,3 (for Italy) mg 9CBC / day. Furthermore, based on our calculations, large parts, i.e. ∼ 2/3rd of the population are low, even too low (<1.1 mg/day), in daily vitamin A5 intake. In addition to the importance of nudging the population towards a regrettably non-well accepted higher intake of fruits and vegetables, an additional fortification and supplementation of vitamin A5 could be considered, similar as to other micronutrients that are low a Westernized diet.

## Introduction

A new vitamin concept was recently proposed for vitamin A5, as the first novel identified vitamin for 80 years [1]. The active form of this vitamin is formed out of its precursors, 9-*cis*-beta-carotene (9CBC), originating from plant derived food items [2], especially from fruits and vegetables, as well as out of 9-*cis*-13,14-dihydroretinol, originating from animal sources [3]. These two vitamin A5 derivatives are the nutritional relevant precursors of 9-*cis*-13,14-dihydroretinoic acid, the molecule relevant for the endogenous activation of the nuclear hormone receptor, the retinoid X receptors (RXRs) [2].

Historically, it was only after clearly observable vitamin deficiencies were identified in humans that the underlying individual and health-relevant mechanism of action was identified [1]. Later on, further detailed dietary recommendations were finally suggested by national and international authorities, such as the USDA in the US [1] or EFSA for Europe [4]. Traditionally, vitamin intake recommendations were based on studies – human observational or even intervention studies or animal ones – that prevented typical, i.e. vitamin related deficiency symptoms [1, 5]. Today, after 80 years since the discovery of the last vitamin [6], different strategies for the identification and establishment of potential novel vitamin concepts are more appropriate. Such a novel strategy is needed, mostly due to ethical restrictions for deprivation and supplemental dose-escalating studies of perceived important dietary constituents in humans [7] and regarding animal studies in sight of difficulties to transfer findings from animal models to humans, due to different metabolism and body weight [7, 8]. In addition, strategies performed around 100 years ago included randomly supplementing people with diseases in a non-ethical and non-standardized way and are not possibly anymore [9], and have been replaced by extremely work intensive and highly expensive standardized clinical procedures [10]. Targeted supplementation trials that exclude specific food components rich in a specific vitamin in a person’s diets, such as performed earlier for vitamin B3 employing war prisoners [11], are for obvious reasons not ethically possible anymore.

Instead, today, a backwards strategy, starting i) from a unique mechanism of action involving the active vitamin form in blood / tissues and ii) relevant cellular concentrations related to normal physiological metabolism towards iii) considering nutritional precursors from food components and calculation of their daily intake amounts, also accounting for iv) aspects of their bioavailability, are a more feasible and logical strategy [12, 13].

In summary, establishing a new vitamin category today requires different strategies and measures than a century ago, especially using state-of-the-art laboratory experiments for establishing a novel food-dependent mechanism of action with crucial health benefits.

Regarding the large picture for vitamin A5, many of these puzzle pieces have been sorted out [1, 2], including its biological relevance as the ligand for the RXR nuclear receptor and the presence of precursor molecules in the diet. However, some crucial parts along the way of establishing intake recommendations can only be addressed following a logical step-by-step procedure connecting the various aspects of tissue concentrations, bioavailability and bioconversion aspects and occurrence in food items together [1].

In this article, we add another important piece to the vitamin A5 story, based on a calculated daily intake of this vitamin in Westernized countries. Based on these estimations, the nutritional situation of these countries with respect to vitamin A5 supply can be evaluated and we can further stratify which subgroups regarding age, gender, or country, may be potentially low in their daily intake levels of this novel vitamin.

## Materials and methods

### Vitamin A5 / Provitamin A5 levels in food items

First, we summarized, based on published studies and databases [14, 15], the vitamin A5 levels in the form of provitamin A5 (9CBC) in food items, focusing on fruits and vegetables as provitamin A5-rich sources that are also frequently consumed. For this purpose, a review of published peer-reviewed literature was conducted to identify relevant data on vitamin A5 levels in food items [1].

In addition to the EFSA food consumption database [15], one additional study that received particular attention was the EPIC cohort [14], as fruit and vegetable intake was available at relevant sub-categories, i.e. leafy vegetables, cabbages, root vegetables, fruiting vegetables, onions/garlic as well as other vegetables, which was not the case for the EFSA food consumption database [15]. We combined the averaged data of 9CBC concentrations in vegetable and fruit subgroups from Table 1A to calculate a weighted average of 0.2 µg 9CBC / g fruits and 5.2 µg 9CBC / g vegetables (Table 1B).

**Table 1:**
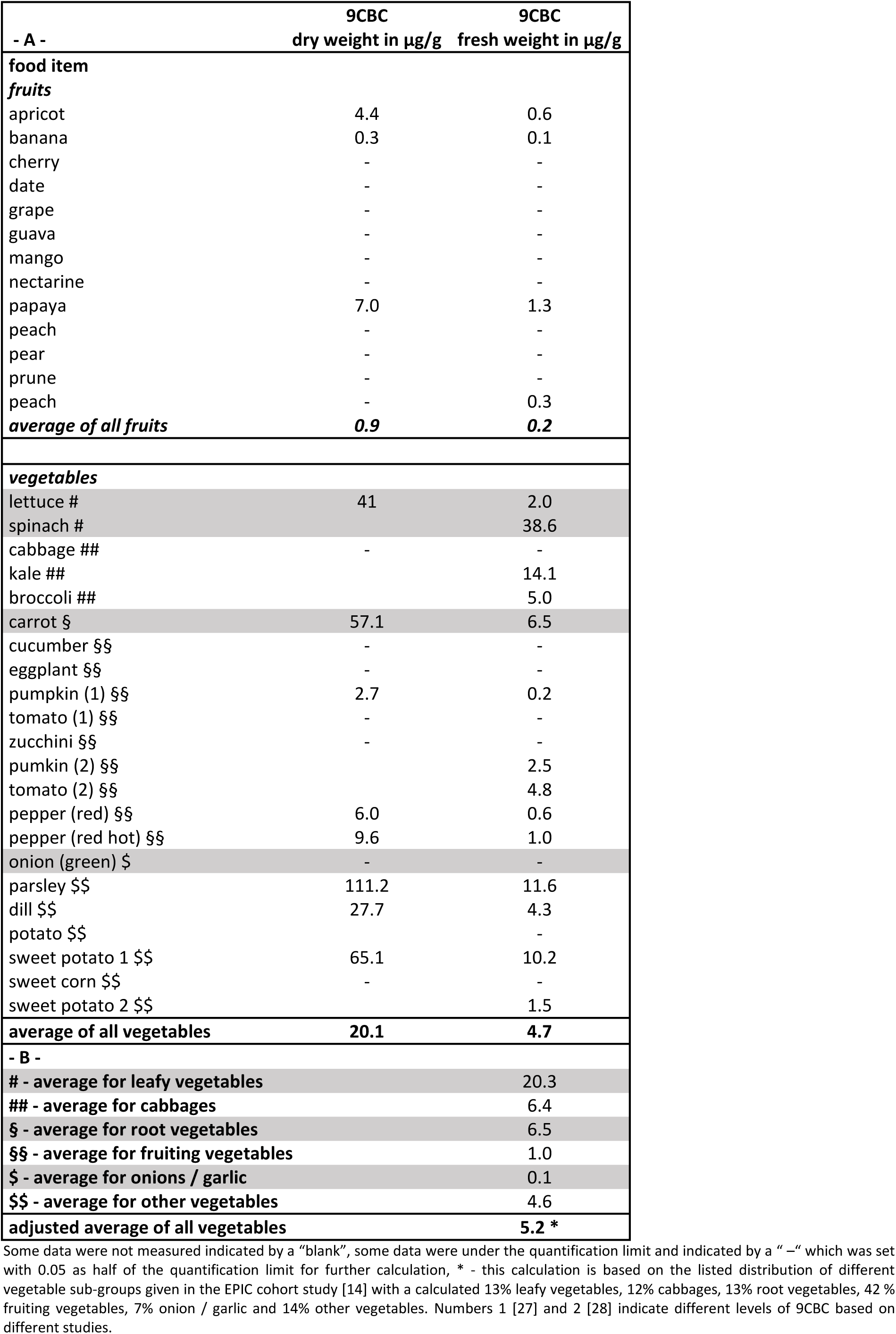
Concentration of 9CBC in µg 9CBC/g food matrix.

### Consumption of fruits and vegetables as well as their subgroups

In a second step, we summarized, based on European food consumption data [15], the average amounts of these food items (i.e. fruits and vegetables) consumed by different populations in individual European countries. For this purpose, due to the large amount of available data and due to the standardized approach by which the data was obtained, we have focused largely on EFSA’s food consumption database summarizing food consumption patterns across member states (displayed in Table 2) [16] and the EPIC cohort based database displayed in Table 3 [14].

**Table 2:** Fruits (F) and vegetables (V) intake (g per day) from the EFSA database of different European countries^Ϯ^.

| Countries | Fruits | Vegetables | F + V | % V | Measurement method and Survey name | Sample size | Year |
| --- | --- | --- | --- | --- | --- | --- | --- |
| Italy | 171 ± 143 | 236 ± 155 | 407 | 58 | Italian national dietary survey on the adult population from 10 up to 74 years old | 726 | 2018 |
| Hungary | 138 ± 139 | 235 ± 166 | 373 | 63 | Hungarian national food consumption survey | 529 | 2018 |
| Serbia | 137 ± 164 | 217 ± 165 | 354 | 62 | Serbian Food Consumption Survey on adults | 1150 | 2019 |
| Luxembourg* | 287 ± 268 | 216 ± 172 | 503 | 43 | ORISCAV-LUX 2 [29] | 1326 | 2017 |
| Latvia | 141 ± 170 | 203 ± 133 | 345 | 59 | Latvian National Dietary Survey | 1080 | 2012 |
| Cyprus | 111 ± 135 | 201 ± 145 | 312 | 65 | National dietary survey of the adult population of Cyprus | 272 | 2014 |
| Greece | 121 ± 164 | 192 ± 143 | 313 | 61 | The EFSA-funded collection of dietary and related data in the general population aged 10-74 years in Greece | 260 | 2014 |
| Portugal | 161 ± 140 | 182 ± 130 | 343 | 53 | National Food, Nutrition, and Physical Activity Survey of the Portuguese general population | 3102 | 2015 |
| Croatia | 136 ± 153 | 174 ± 158 | 310 | 56 | Croatian food consumption survey on adults | 2002 | 2011 |
| Estonia | 226 ± 231 | 168 ± 159 | 394 | 43 | National Dietary Survey among 11-74 years old individuals in Estonia | 2124 | 2013 |
| Montenegro | 120 ± 110 | 163 ± 121 | 326 | 50 | Montenegrin National Dietary Survey on the general population | 697 | 2017 |
| Netherlands | 115 ± 128 | 163 ± 117 | 279 | 59 | Dutch National Food Consumption Survey 2012-2016 (DNFCS) | 1487 | 2012 |
| Romania | 163 ± 186 | 154 ± 110 | 317 | 49 | Romanian national food consumption survey for adolescents, adults, and elderly | 740 | 2019 |
| Slovenia | 158 ± 151 | 153 ± 105 | 311 | 50 | Slovenian national food consumption survey | 385 | 2017 |
| Ireland | 91 ± 95 | 151 ± 84 | 242 | 62 | North/South Ireland Food Consumption Survey | 958 | 1997 |
| Bosnia and Herzegovina | 130 ± 144 | 149 ± 122 | 279 | 53 | Bosnia-Herzegovinian Dietary Survey of adolescents, adults, and pregnant women | 850 | 2017 |
| Spain | 155 ± 129 | 144 ± 102 | 299 | 48 | Spanish National dietary survey in adults, elderly, and pregnant women | 536 | 2013 |
| France | 133 ± 131 | 144 ± 90 | 277 | 52 | Individual and national study on food consumption 2 | 2276 | 2007 |
| Finland | 171 ± 178 | 126 ± 102 | 297 | 43 | National Findiet Surveys | 1575 | 2007 |
| Sweden | 91 ± 104 | 126 ± 99 | 218 | 59 | National Diet and Nutrition Survey - Years 1-3 | 1266 | 2008 |
| Belgium | 118 ± 122 | 125 ± 100 | 244 | 52 | Diet National 2004 | 1292 | 2004 |
| Czech Republic | 124 ± 123 | 117 ± 91 | 241 | 49 | Czech National Food Consumption Survey | 1666 | 2003 |
| Germany | 165 ± 175 | 95 ± 94 | 260 | 37 | National Nutrition Survey II | 10.419 | 2007 |
| Austria | 162 ± 153 | 90 ± 96 | 252 | 36 | Austrian Study on Nutritional Status 2010-12 - Adults | 308 | 2010 |
| European average ± STD | 147 ± 42 | 164 ± 41 | 312 ± 64 | 53 ± 8 | Calculated sum of the 24 listed surveys | 37026 |  |

Countries were sorted from top to bottom, based on the average vegetable intake.
† European Food consumption data [16]). Age range from 18-64 years.
\* median and interquartile range

**Table 3:** Summarised data from the EPIC cohort based on the data from the year 2002 [14].

| men |  | fruits (F) | vegetables (V) | F + V | leafy V | % V / F+V | % LV / V |
| --- | --- | --- | --- | --- | --- | --- | --- |
| Country | n | mean | mean |  | mean |  |  |
| Greece | 1312 | 273 | 270 | 543 | 30 | 50 | 11 |
| Spain | 1777 | 372 | 224 | 596 | 44 | 38 | 19 |
| Italy | 1444 | 403 | 211 | 614 | 35 | 34 | 17 |
| Germany | 2268 | 207 | 160 | 368 | 14 | 44 | 9 |
| Netherlands | 1024 | 168 | 137 | 305 | 28 | 45 | 20 |
| UK * | 518 | 206 | 193 | 400 | 10 | 48 | 5 |
| Denmark | 1923 | 160 | 141 | 301 | 10 | 47 | 7 |
| Sweden | 2765 | 122 | 112 | 234 | 9 | 48 | 8 |
| sum # / av ## | 13031 | 239 | 181 | 420 | 23 | 44 | 12 |
| STD |  | 102 | 53 | 146 | 13 | 5 | 6 |
| women |  | fruits (F) | vegetables (V) | F + V | leafy V | % V / F+V | % LV / V |
| Country | n | mean | mean |  | mean |  |  |
| Greece | 1374 | 242 | 207 | 449 | 29 | 46 | 14 |
| Spain | 1443 | 355 | 196 | 551 | 34 | 36 | 17 |
| Italy | 2512 | 343 | 181 | 524 | 28 | 34 | 16 |
| Germany | 2150 | 236 | 166 | 403 | 17 | 41 | 10 |
| Netherlands | 2960 | 192 | 130 | 322 | 23 | 40 | 18 |
| UK * | 768 | 223 | 192 | 415 | 15 | 46 | 8 |
| Denmark | 1995 | 206 | 150 | 356 | 11 | 42 | 7 |
| Sweden | 3285 | 164 | 127 | 290 | 11 | 44 | 9 |
| France ** | 4639 | 251 | 226 | 476 | 45 | 47 | 20 |
| Norway ** | 1798 | 168 | 125 | 293 | 7 | 43 | 5 |
| sum # / av ## | 22924 | 238 | 170 | 408 | 22 | 42 | 12 |
| STD |  | 66 | 36 | 93 | 12 | 4 | 5 |
| women and men |  | fruits (F) | vegetables (V) | F + V | leafy V | % V / F+V | % LV / V |
| Country | n | mean | mean |  | mean |  |  |
| Greece | 2686 | 257 | 238 | 496 | 29 | 48 | 12 |
| Spain | 3220 | 363 | 210 | 573 | 39 | 37 | 18 |
| Italy | 3956 | 373 | 196 | 569 | 32 | 34 | 16 |
| UK * | 1286 | 215 | 193 | 407 | 13 | 47 | 6 |
| Germany | 4418 | 222 | 163 | 385 | 15 | 42 | 9 |
| Denmark | 3918 | 183 | 145 | 328 | 10 | 44 | 7 |
| Netherlands | 3984 | 180 | 133 | 313 | 25 | 43 | 19 |
| Sweden | 6050 | 143 | 119 | 262 | 10 | 46 | 8 |
| sum # / av ## | 29518 | 242 | 175 | 417 | 22 | 43 | 12 |
| STD |  | 85 | 41 | 118 | 11 | 5 | 5 |

The countries are sorted from top to bottom, based on the vegetable intake of women and men, n – number of included persons, av – average, \* just health aware people were included, \*\* just woman were examined for this country, # - the sum is relevant for the number (n) of included persons, ## - average of fruits intake, vegetable intake, F + V intake, leafy V intake, % V / F+V and % LV / V.

The EFSA food consumption database provided detailed data on food intake patterns, including frequency and quantity of fruit and vegetable consumption, across various demographic groups (age and gender based). When several populations or studies were available for a European country, the arithmetic mean per each study was first calculated. Thereafter, the average consumption per country level was calculated, which was then termed “mean of means”.

In addition to food intake data, information was also collected on fruit and vegetable intake recommendations per country, in order to compare estimated intakes with the respective countries’ food based dietary guidelines.

### Calculation of provitamin A5 intake

As a third step, Further we calculated the intake of 9CBC by means of the more detailed EPIC cohort data, by combining the average 9CBC contents from Table 1B with the food intake data from EPIC (Table 3) per country. Furthermore, we calculated, based on these estimations and the previous intake estimates the amount of provitamin A5 consumed per day based in each European nation (Figure 3 and 4).

### Statistical Analysis

Descriptive statistics, including mean and standard deviation (SD), were computed to summarize the average daily intake levels of provitamin A5 across different population groups. Frequency distributions were also examined to identify patterns of vitamin A5 intake within each population.

## Results

### Concentration of vitamin A5 in the form of provitamin A5 in individual food items

A limited number of publications exists reporting concentrations of vitamin A5 / 9CBC, expressed as provitamin A5, i.e. 9CBC, in fruits and vegetables (Table 1). The average concentration of 9CBC in fresh fruits was found to be 0.2 µg 9CBC / g fruits and the estimated concentration in vegetables was 4.7 µg 9CBC / g vegetables, while a weighted average estimation, considering different intake amounts of individual vegetable subgroups, resulted in an average of 5.2 µg 9CBC / g vegetables, which was used for all further calculations.

When comparing different sub-groups of vegetables, the highest concentrations appeared to be present in leafy vegetables, followed by root vegetables and cabbages. It should be noted that fruit juices, for lack of data regarding 9CBC, were not included in this evaluation.

Data regarding vitamin A5 intake in the form of 9CDHROL and 9CDHROL-esters from animal based products were not reported, as there are currently no data available and concentrations have been reported to be very low [2].

### Summary of current intake recommendations for fruits and vegetables

### Daily intake of fruit and vegetables in Europe

#### A. Based on the EFSA food consumption database

Data from 24 countries, 23 based on EFSA’s food consumption database, encompassing a total of 26.600 participants aged 18-64 years were included in this investigation. As shown in figure 1 and table 2, fruit intake ranged from 91 g / day (Sweden and Ireland) to 287 g/day (Luxembourg) with an average of 147 g / day. Vegetable intake was reported lowest in Austria (90 g / day) and highest in Italy (236 g / day) and an average of 164 g / day. This translates into a highest combined intake of fruits and vegetables with 503 g / day in Luxembourg, lowest in Sweden with 218 g / day and an average of 312 g / day.

**Figure 1:**
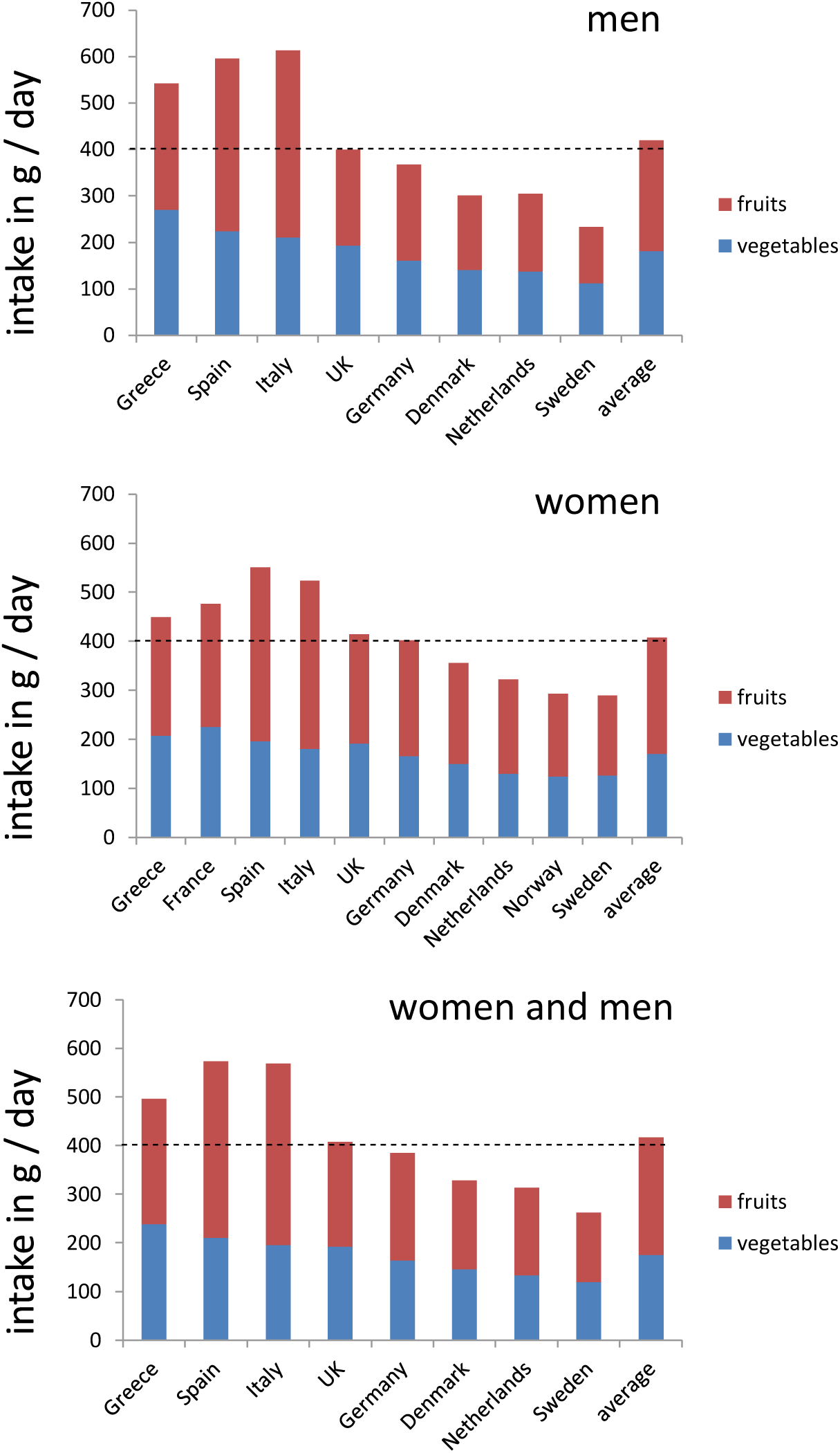
Schematic summary of a dietary pattern focusing on fruit and vegetable intakes in different European countries. *The dashed line indicates the recommended “5 A DAY” recommendation of 5 portions of 80 g fruits and / or vegetables for daily intake*.

Most countries did not even reach the minimum WHO recommended 400 g fruits and vegetables / day: According to EFSA’s food consumption database, out of the 23 countries listed in table 2, with only Italy and Luxembourg reaching the recommended 400 g of fruits and vegetables / day (i.e. 5 portions per day) target. Especially Southern European countries such as Spain, Greece, Portugal, Italy and Cyprus showed generally higher combined fruit and vegetable intakes compared to Northern European countries such as Ireland, Denmark, Sweden, or Finland.

#### B. The EPIC cohort

When comparing findings by EFSA to the EPIC cohort, in which data was generally also obtained by 24 h recalls (though not by multiple ones), similar tendencies were found for fruit and vegetable intakes with a higher average of 417 g / day in comparison to 312 g / day in the EFSA database. The average vegetable intake in the EPIC database was 175 g / day and comparable to the 164 g / day in the EFSA database. The average fruit intake was 242 g / day in the EPIC cohort while the EFSA values were much lower with 147 g / day.

Highest intake of fruits / vegetables per capita (Table 3) were found comparable to the EFSA database in the Southern European Nations like Spain (573 g / day), followed by Italy (569 g / day) and Greece (496 g / day), while lowest once were mainly determined also comparable in Northern European countries like Sweden (262 g / day) and the Netherlands (313 g / day). With regard to the recommended 400 g of fruit and vegetables per day, only 4 out of 8 countries (Table 3 and Figure 1), for which data were available, for both men and women met this intake recommendation.

### Calculation of individual intake of vegetable subgroups

Regarding vegetable subgroups as determined based on the EPIC cohort study, a total of 22 g leafy vegetables (men and women combined, out of 175 g total vegetables) were consumed, based on data from 8 countries reporting both data of men and women (Table 3).

Regarding fruits, an average of 242 g / day were consumed. Southern countries such as Italy, Spain and Greece showed consistently higher intakes for vegetables, including leafy ones and fruits compared to Northern European countries such as Denmark, the Netherlands, Germany and Sweden. An exception was noted for cabbages, which did not follow this trend, as high concentrations were consumed in the UK, Denmark and Germany. When looking at subgroups in man or woman, no remarkable overall differences were apparent for the intake of total vegetables, leafy vegetables, and individual subgroups (Figure 2).

**Figure 2:**
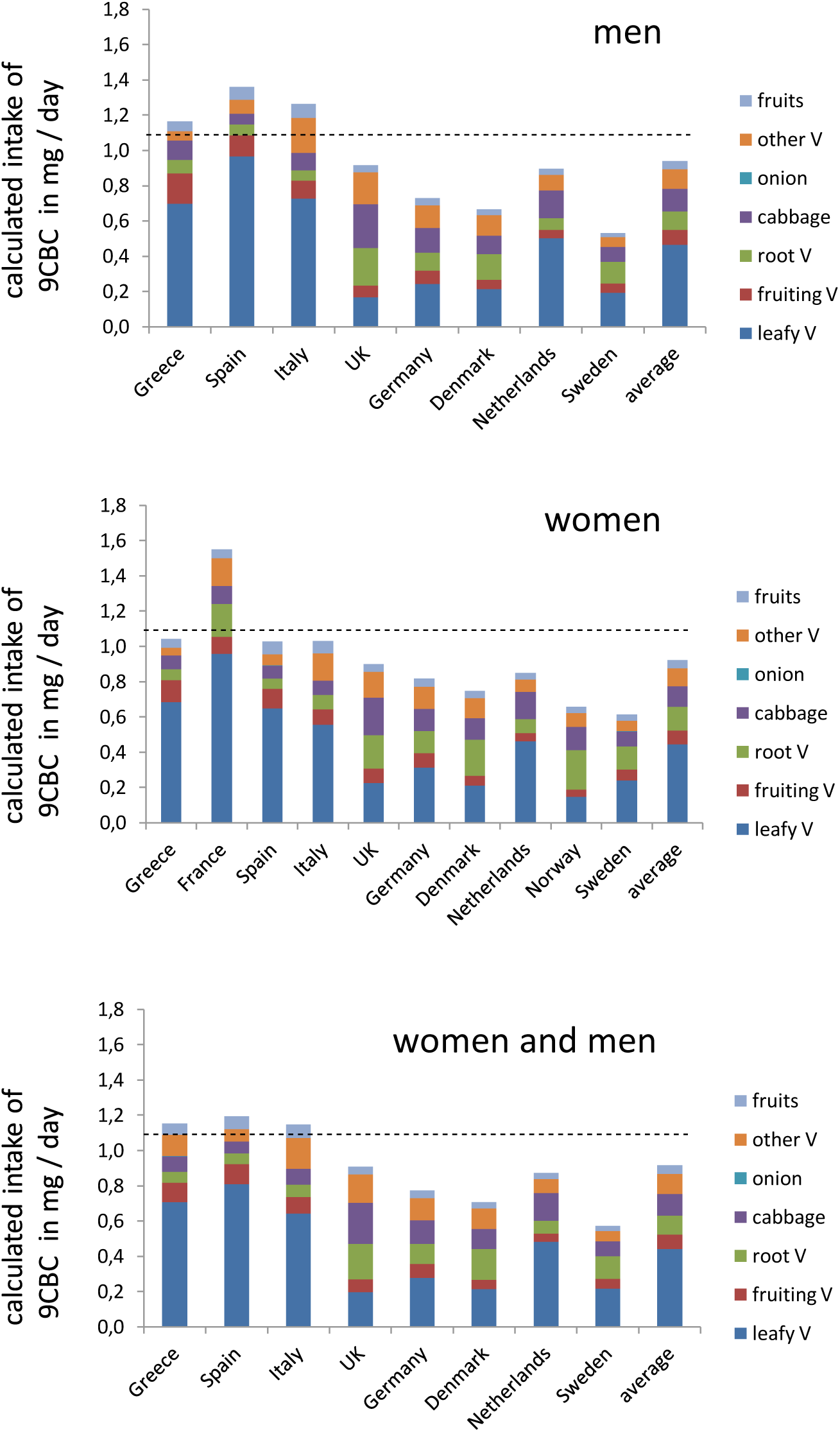
Schematic summary of the calculated vitamin A5 intake levels in different European countries. Used abbreviations; V - vegetables

### Estimating the daily intake of provitamin A5

Based on the EPIC cohort about half of the countries would fall below the earlier proposed 1.1 mg / day intake of provitamin A5 (Figure 2 and 3). Only the Southern European countries, due to their generally higher intake of fruits and vegetables (Tables 2-3 and Figure 2) were estimated to reach the 1.1 mg / day target, while Northern and Central European countries had lower intakes as 0.5 mg / day. Employing data from EFASs Food Consumption database, and also assuming an approx. content of 9CBC of 0.2 µg / g for fruits and 5.2 µg / g for vegetables (based on Table 1B) based on this cruder estimate, only few countries like Italy, Hungary, Luxembourg and Serbia reached the 1.1 mg / day 9CBC target while the majority of 19 countries did not reach the suggested 1,1 mg 9CBC intake / day. The European average was comparable with 0.9 mg 9CBC / day in the EPIC cohort and 0.9 mg 9CBC in the EFSA cohort (Figure 3).

**Figure 3:**
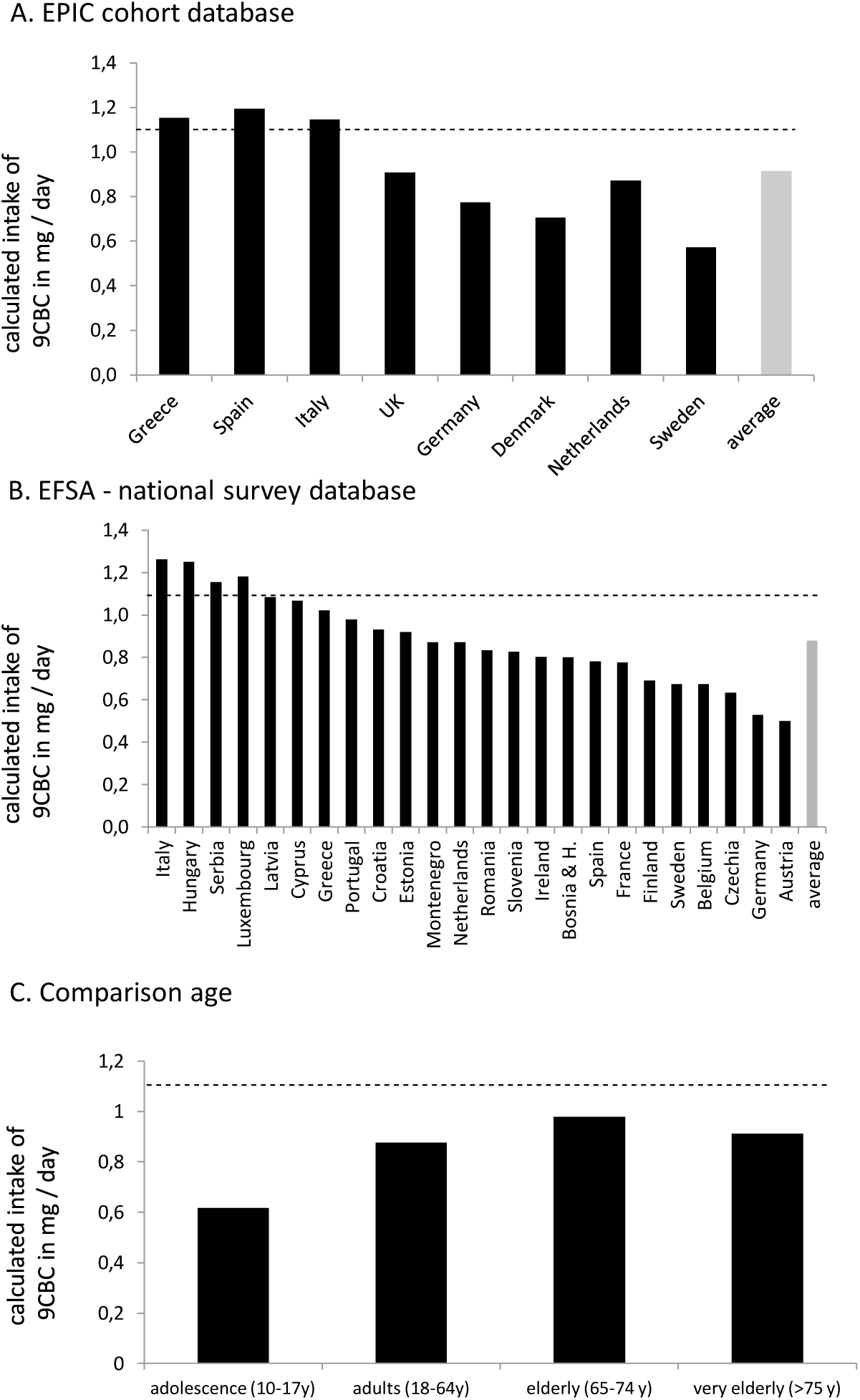
Calculated individual daily 9CBC intake levels based from the data of the a) the EPIC cohort [14], b) summarized national surveys and c) age depending intake, both based on EFSAs Food Consumption database [16] : All estimated values are based on average values for 9CBC in fruits (0.2 µg/g) and vegetables (5.2 µg/g). *The dashed line indicates the recommended intake of 1.1 mg 9CBC (provitamin A5) /day* [1].

Due to their high content of 9CBC combined with their relatively frequent consumption, leafy vegetables were the vegetable subgroup most largely contributing to total provitamin A5 intake (contributing to often >50% of total 9CBC intake), followed by root vegetables, cabbages and other vegetables (Figure 2). Fruits contributed, due to their low content of 9CBC, very little to overall intake, below 5% (Figure 2). Slight differences between men and women were noted, with women showing slightly lower average intakes of 9CBC.

When looking at 9CBC intake by age in the EFSA cohort (Figure 3C), the differences between the age groups appeared to be small, with the highest intakes in elderly adults aged 65-74 years. This is somewhat confirmed by EFSA data [15], as fruit and especially vegetable consumption is highest also for the elderly compared to adolescents and the elderly.

### Comparison of provitamin A5 intake in persons consuming low, medium and high amounts of fruits and vegetables (Figure 4)

**Figure 4:**
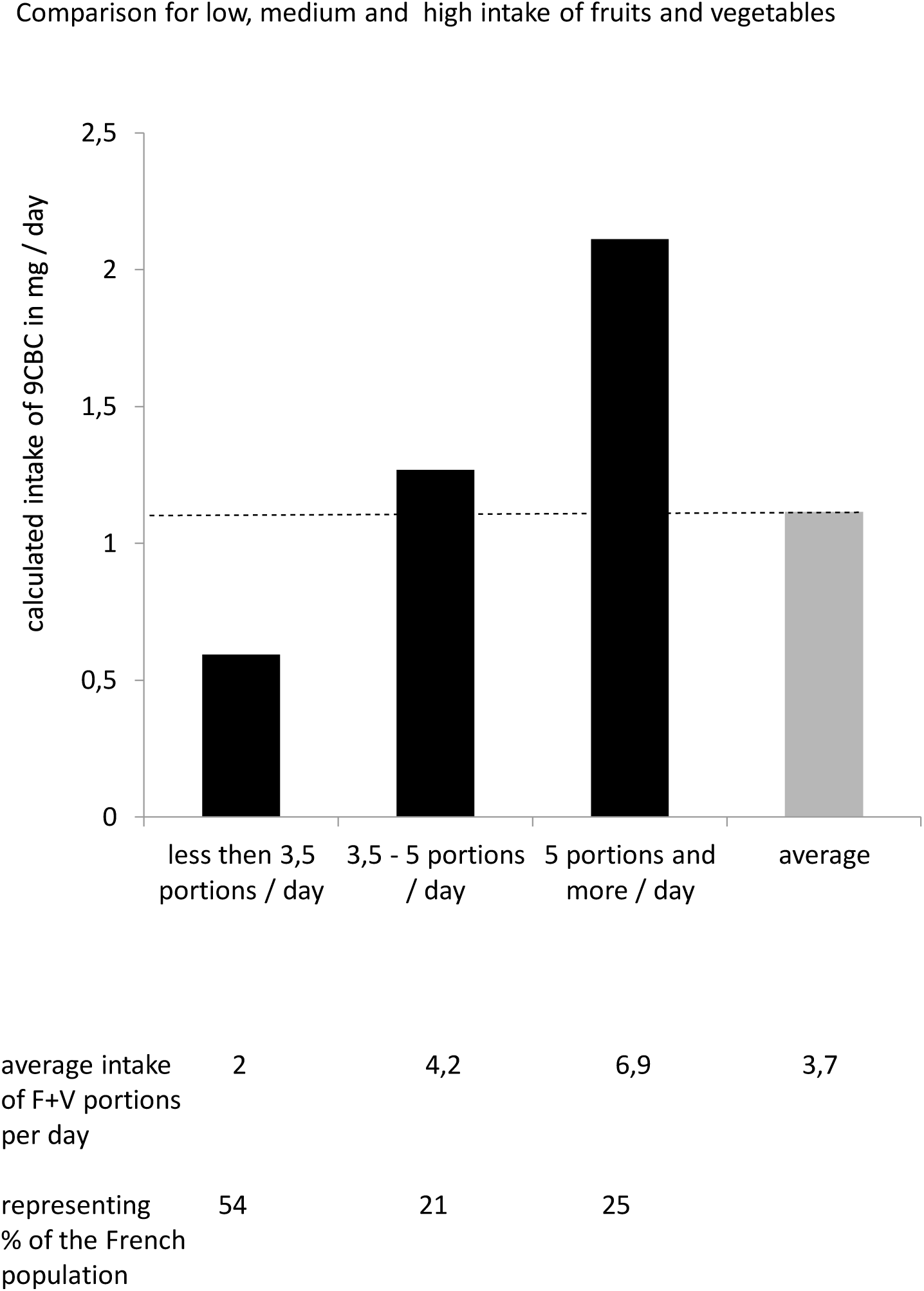
Calculated daily 9CBC intake levels of individuals with low, medium and high daily average fruit and vegetable intake from a French survey [14] with an average F+V intake of 295 g F+V / day. *The dashed line indicates the recommended intake of 1.1 mg 9CBC (provitamin A5) /day* [1].

Based on data from a study just performed in France [17], the majority (>50%) consumed less than 3.5 portions fruits and vegetables / day with an average of 2 portions, reach in consequence an average intake of only 0.6 mg provitamin A5 / day. Furthermore, only 21% of the French population consumed between 3,5 and 5 portions of fruits and vegetables per day, based on an average of 4,2 portions of fruits and vegetables / day and consequently an estimated provitamin A5 intake of 1,3 mg 9CBC / day [18]. Only 25 % of the French population consumed sufficient fruits and vegetables with 5 and more portions / day and an estimated provitamin A5 intake of 2,1 mg 9CBC / day [18, 19]. In consequence, this means that ∼64% (∼2/3^rd^) of the French population consume to few fruits and vegetables per day and in consequence also a tow low daily intake of provitamin A5.

## Discussion

In this article we present, for the first time, an estimation of vitamin A5 intake in various European countries. This compound has been identified and categorized as a novel vitamin and / or an independent subcategory of vitamin A and an intake estimate across various European populations was required. Toward this end, we calculated vitamin A5 intakes independently from vitamin A(1) from the background diet. An average intake of vitamin A5 in the adult population ranging from 0.5 to 1.3 mg per day was found, with a tendency for higher intakes in Southern European countries. It was also estimated that approx. 2/3 of the population would consume less than the proposed recommended intake of 1.1 mg provitamin A5 / day.

Comparable to other vitamins, we make use of available data from cohort studies, in our case from the EPIC [14] and EFSA cohorts [15], allowing to calculate the daily intake specified for several European countries, sex and partly age groups, which are currently available in these databases. As a first step, based on available data, we calculated the average intake of vitamin A5 in food categories that are rich in vitamin A5. Here we focused on fruits and vegetables, where provitamin A5, i.e. 9-*cis*- β-carotene, is mainly present. The levels of 9-*cis*-13,14-dihydroretinol, as well as its esters in the human food matrix have thus far either not been determined or are deemed very low [3], suggesting that the main intake of vitamin A5 is via plant foods rich in provitamin A5 [20]. At first, an average provitamin A5 concentration was calculated, with 0.2 µg provitamin A5 / day for fruits and 5.2 µg provitamin A5 / day for vegetables, which were then employed for further calculations. This was required, as most databases do not contain further details on the consumption of individual fruits and vegetable subgroups.

The highest daily intake amounts of fruits and vegetables was determined in Southern European countries such as Greece, Spain and Italy, and lowest in Northern European countries including Sweden, the Netherlands and Denmark. Remarkable is that the large majority of Europeans was consuming less than the suggested “5 A DAY”-intake recommendations of 5 portions of each 80 g fruits and vegetables by national and international food organizations [14, 15, 21]. However, this has been remarked in earlier consumption surveys in Europe [14].

Regarding the best sources of provitamin A5 per weight, the main source of provitamin A5 intake originates from the vegetable sub-group leafy vegetables, with 20.3 µg provitamin A5 / g food matrix, followed by cabbages (6.4 µg/g) and root vegetables (6.4 µg/g), with low concentrations in fruiting vegetables (1.0 µg/g) and onions / garlic (0.1 µg/g). The European country-specific daily intake amount of these leafy vegetables was highest in Southern European countries such as Greece, Spain and Italy and lowest in Northern / Central European countries including UK, Norway, Denmark and Sweden. This is in line with studies reporting higher fruit and vegetable intake in general in Southern European countries [14].

Based on the fruit and vegetable intake amounts from the EPIC and the EFSA database [14, 15], summarizing 65,544 individuals, we calculated for 27 European countries country specific daily vitamin A5 intake amounts with an average of 0.9 mg provitamin A5 / day, which is lower than the previously suggested recommendation of 1.1 mg provitamin A5 / day, calculated by two independent methods [1]. These estimated average intake amounts of provitamin A5 on average of European countries sound moderate / fair compared to the suggested intake amounts. While when stratifying for different countries, we observed that the average in the non-Southern European countries such as especially Austria (0.5 mg/day), Germany (0.5 mg/day), Czech Republic (0.6 mg/day), Sweden (0.7 mg/day EFSA data / 0.6 mg/day EPIC data) were clearly below the suggested intake amounts of 1.1 mg provitamin A5 / day. Based on the estimations of daily provitamin A5 intake from the 27 European countries, only 4 out of 24 countries for the EFSA database calculations and 3 out of 8 from the EPIC database calculations reach sufficient high daily provitamin A5 intake levels on average, while the large majority of the European countries 3 (based on the EPIC database) and 8 (based on the EFSA database) being even below 0.8 mg provitamin A5 / day on average and thereby far below the suggested daily intake amounts of provitamin A5. This means that the large majority of European countries on average have a too low daily intake of daily vitamin A5. However, it also needs to be specified that both the intake recommendation as well as the calculated intake are based on average values for adults, and due to the limited amount of data available. Further studies should aim to determine detailed estimations such as allowing estimating an intake that would cover over 97% of the population’s need, such as necessary to determine the population reference intake (PRI).

Further based on evaluations in France [17] it was calculated that 54% of the French population eat less than 3.5 daily portions of fruits and vegetables and a further considerable fraction (21 %) consume between 3.5 – 5 portions, which means that ca. 64% of the French population are below the suggested “5 A DAY” recommendations. This means as a consequence that ∼2/3 of the French population, as well as transferable to many other European populations are below the suggested daily intake levels of vitamin A5 which were based on the “5 A DAY”-concept.

In summary, due to the low intake of fruits and vegetables [22], especially vegetables [14] and with special focus on low intake of leafy vegetables [14], there is an estimated low, even too low, daily dietary intake of vitamin A5 present in Europeans. This data is likely relevant for other rather Western-lifestyle based societies following western based dietary patterns, which includes large populations in many developing countries [1, 23]. We recommend, based on these calculated parameters that, in order to improve dietary patterns and assure also sufficient intake of vitamin A5 that a) novel dietary recommendations should focus mainly on a higher daily dietary intake of vegetables and especially leafy vegetables and b) consider additional food fortification with vitamin A5 and / or dietary supplements to meet the suggested daily dietary intake recommendations for vitamin A5 to avoid a potential vitamin A5 deficiency [20].

Whether a low daily intake of vitamin A5 is associated with mental problems, which are prevalent in our Western hemisphere [20] is currently merely predicted and requires further attention based on European-wide epidemiological databases with sufficient details for specific European countries regarding dietary intake and mental health issues. Not surprisingly, in Southern European countries like Greece, Italy and Croatia with a high average estimated vitamin A5 intakes, the mental problems appear to have a lower prevalence, while in Northern European countries like the Netherlands, Finland, Denmark, Sweden and Germany these mental problems are reported to have a high prevalence [24]. A direct correlation and causal connection must still be elaborated and evaluated in larger databases, together with deeper functional mechanistic evaluations. Furthermore, described and suggested long-term or medium-term vitamin A5-deficiencies such as poor mental health and high incidence of numerous neurological diseases should be evaluated with respect to nutritional or functional vitamin A5-signalling pathways [20]. While poor mental health is described to be based on low daily intake of fruits and vegetables, especially vegetables [25] and with a special focus on leafy vegetables [26]. A detailed clinical mechanistic step-by-step approach how fruits and vegetables, with a focus on vegetables and leafy vegetables rich in vitamin A5, transmit their health-promoting and –protecting activity, via the retinoid X receptor, was summarized in a recent article [1], which is based on dozens of mechanistic studies performed using laboratory models as a base.

In this study, a more detailed and data-based intake of vitamin A5 in several European countries was attempted and results were compared against a first recommendation for vitamin A5 intake. The calculated data were evaluated in an ideal case dietary scenario as well as in a reality-based scenario [1], showing that large parts of the European society may be too low in daily intake of vitamin A5.

## Data Availability

All data produced in the present study are available upon reasonable request to the authors

## Author contributions

RR was responsible for the idea and outline and writing of the article, FV did export and calculate food consumption data, TB was involved in writing the article and critically reviewing the concept.

## Conflict of interest

RR is CEO and shareholder of CISCAREX UG.

## Notes

### Funding Statement

This study did not receive any funding

### Author Declarations

All mentrioned in publications listed.

